# Detecting HRD in whole-genome and whole-exome sequenced breast and ovarian cancers

**DOI:** 10.1101/2024.07.14.24310383

**Authors:** Ammal Abbasi, Christopher D. Steele, Erik N. Bergstrom, Azhar Khandekar, Akanksha Farswan, Rana R. Mckay, Nischalan Pillay, Ludmil B. Alexandrov

## Abstract

Breast and ovarian cancers harboring homologous recombination deficiency (HRD) are sensitive to PARP inhibitors and platinum chemotherapy. Conventionally, detecting HRD involves screening for defects in *BRCA1*, *BRCA2*, and other relevant genes. Recent analyses have shown that HRD cancers exhibit characteristic mutational patterns due to the activities of HRD-associated mutational signatures. At least three machine learning tools exist for detecting HRD based on mutational patterns. Here, using sequencing data from 1,043 breast and 182 ovarian cancers, we trained Homologous Recombination Proficiency Profiler (HRProfiler), a machine learning method for detecting HRD using six mutational features. HRProfiler’s performance is assessed against prior approaches using additional independent datasets of 417 breast and 115 ovarian cancers, including retrospective data from a clinical trial involving patients treated with PARP inhibitors. Our results demonstrate that HRProfiler is the only tool that robustly and consistently predicts clinical response from whole-exome sequenced breast and ovarian cancers.

**SIGNIFICANCE:** HRProfiler is a novel machine learning approach that harnesses only six mutational features to detect clinically useful HRD from both whole-genome and whole-exome sequenced breast and ovarian cancers. Our results provide a practical way for detecting HRD and caution against using individual HRD-associated mutational signatures as clinical biomarkers.

## INTRODUCTION

Repair of DNA double strand breaks by homologous recombination (HR) is an essential cellular mechanism for maintaining genomic stability and for preventing tumorigenesis (1). Prior studies have elucidated key genes in the HR pathway, including, *BRCA1*, *BRCA2, RAD51*, and *PALB2*, that commonly have pathogenic germline variants and/or somatic mutations in breast and ovarian cancers (1). Defects in HR genes can disable the HR repair pathway making cells vulnerable to double strand breaks and, thus, provide a treatment opportunity. Specifically, patients with cancers harboring defective HR repair are sensitive to both poly (ADP-ribose) polymerase inhibitors (PARPi) and to platinum chemotherapy (2,3).

Conventional stratification of HR deficient (HRD) and HR proficient (HRP) cancers involves screening for canonical genomic markers, including pathogenic germline variants and somatic copy number alterations in HR genes (4–6). Previous experimental studies (7) and genomics analyses (8) have also revealed that HRD cells exhibit characteristic patterns of somatic mutations due to the activities of HRD-associated mutational processes. Currently, there are at least seven mutational signatures that have been putatively associated with and/or utilized to detect HRD: *(i)* single base substitution (SBS) signatures SBS3 and SBS8 both characterized by generally flat, yet distinct, profiles (9); *(ii)* genomic rearrangement signatures RS3 and RS5 reflecting non-clustered tandem duplications and deletion, respectively (10); *(iii)* small insertions and deletions (ID) signatures ID6 and ID8, predominately encompassing indels at microhomologies (11); and *(iv)* copy number (CN) signature CN17 characterized by large tandem duplications (12).

At least three machine learning approaches have also been developed to capture HR deficient cancers by examining the patterns of somatic mutations found in cancer genomes: HRDetect (13), CHORD (14), and SigMA (15). HRDetect uses signatures SBS3, SBS8, RS3, RS5, and indels at microhomologies corresponding to ID6 and ID8 to detect HRD in breast cancers (13). CHORD is an alternative pan-cancer HRD prediction tool that does not rely on mutational signatures, but it rather uses 29 mutational features directly observed in cancer genomes (14). CHORD is more computationally efficient and prior studies have shown that it has an almost identical performance to the one of HRDetect (13). However, both CHORD and HRDetect use HRD-specific patterns of genomic rearrangements that can be only reliably detected from whole-genome sequencing (WGS) data (13,14). By excluding genomic rearrangements, HRDetect can also be applied to whole-exome sequencing (WES) data, albeit, with significantly diminished performance (13). Conversely, CHORD’s implementation does not allow utilizing WES cancers. In contrast to CHORD and HRDetect, SigMA was developed to exclusively detect HRD-associated signature SBS3 from whole-genome, whole-exome, and targeted gene panel sequencing data with SigMA’s focus being on panel sequencing data (15). Nevertheless, to be applied to a sample, SigMA requires at least five somatic mutations within the examined cancer (15). Based on Memorial Sloan Kettering Cancer Center’s Integrated Mutation Profiling of Actionable Cancer Targets (MSK-IMPACT) data (16), this limits SigMA’s applicability to approximately 37% of breast and ovarian cancers profiled with MSK-IMPACT targeted gene panel.

In this manuscript, we perform retrospective analyses to evaluate the clinical utility of canonical gene-based biomarkers, HRD-associated mutational signatures, and machine learning approaches to detect treatment sensitive breast and ovarian cancers. While the presence of individual HRD-associated mutational signatures are generally ineffective in detecting clinical response, existing machine learning tools can capture treatment sensitivity in WGS cancers but not in WES cancers. To address this limitation, we developed Homologous Recombination Proficiency Profiler (HRProfiler), a machine learning method that harnesses only six mutational features for detecting clinically actionable HRD from both whole-genome and whole-exome sequenced breast and ovarian cancers. Our findings offer a pragmatic approach to detect HRD in WES cancers and underscore the importance of exercising caution when considering individual HRD-associated mutational signatures as clinical biomarkers.

## RESULTS

### Feature engineering and model training of HRProfiler

To determine the set of robust HRD-associated mutational patterns that can be detected using WGS and WES cancers, we identified significantly enriched mutation types specific to somatic SBSs (9), IDs (11), and CNs (12). In particular, using previously developed schemas (9,11,12), we compared the types of somatic mutations enriched in HRD or HRP cancers. Comparisons were performed for whole-genome sequenced breast cancers using a subset of the Sanger Institute’s 560 breast cancer genomes cohort (10) (Sanger-WGS-Breast; **Fig. 1*a***) as well as for whole-exome sequenced breast cancers using a subset of TCGA’s breast cancer cohort (17) (TCGA-WES-Breast; **Fig. 1*b***). As previously done (13,14,18) patients were classified as HRD based on a combination of their genomic instability and the presence of pathogenic germline variants, somatic mutations, or methylation of *BRCA1* or *BRCA2*. Feature engineering and the subsequent training of HRProfiler was performed only on the designated training datasets (**Supplementary Fig. S1**).

**Figure 1:**
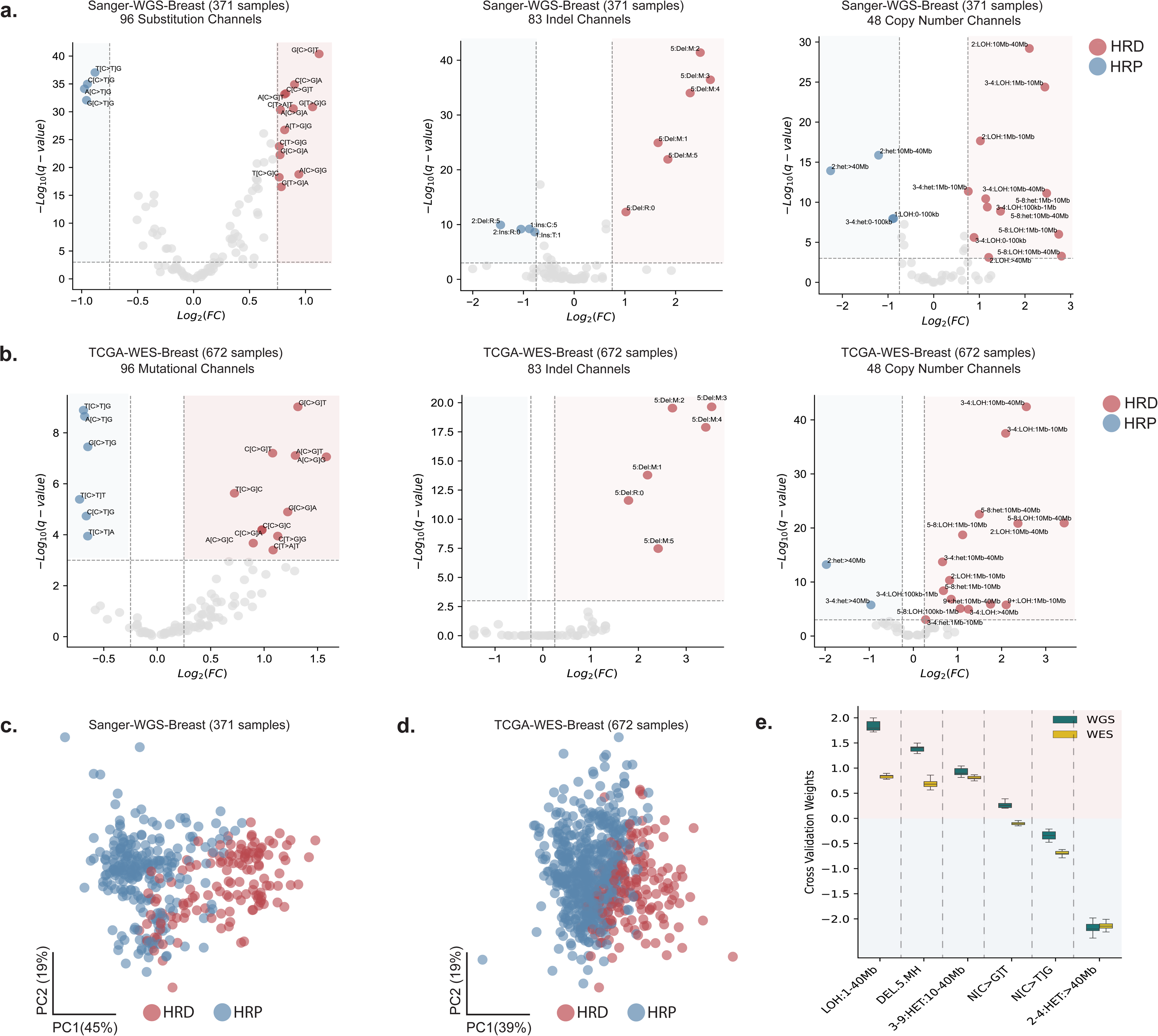
Feature engineering to identify significantly enriched somatic mutational features across HRD and HRP breast cancers. ***(a-b)*** Volcano plots with log_2_ fold change (FC) enrichments across the average proportions of somatic mutations for 96 substitution, 83 indel, and 48 copy number mutational channels between homologous recombination deficient (HRD) and homologous recombination proficient (HRP) cancers for 371 Sanger-WGS-Breast *(a)* and 672 TCGA-WES-Breast samples *(b)*. Channels with an absolute FC greater than 0.75 for WGS and 0.25 for WES, and a −log_10_ FDR adjusted p-value greater than 3 are colored. Channels colored in red are enriched in HRD samples, while channels highlighted in blue are enriched in HRP samples. ***(c-d)*** Principal component (PC) analysis highlights the relevance of the features derived from the significant channels in *(a-b)* by separating HRD from HRP samples across the 371 Sanger-WGS-Breast *(c)* and 672 TCGA-WES-Breast cohorts *(d)*. ***(e)*** The average 10-fold cross validation weights of the six features derived from the WGS and WES breast training datasets using a linear-kernel support vector machine. Positive weights reflect features predictive for HRD samples, while negative weights correspond to features predictive for HRP samples.

At the SBS resolution, we observed a striking enrichment of C:G>T:A single base substitutions at 5’-Np<u>C</u>pG-3’ context (mutated based underlined; N reflects any base) in HRP samples (**Fig. 1*a-b***). This suggests that a relatively large proportion of mutations in HRP samples are C:G>T:A transitions at CpG sites when compared to HRD samples. Conversely, HRD samples were enriched for C:G>G:C single base substitutions at 5’-Np<u>C</u>pT-3’ context. At the indel resolution, we observed an enrichment of deletions spanning at least 5 base pairs (bp) with flanking microhomology sequences across HRD samples (**Fig. 1*a-b***). These mutations are known to arise from the erroneous activities of the microhomology-mediated end joining or the single strand annealing DNA repair pathways in the absence of a functional HR pathway (19). At the copy number resolution, Loss of Heterozygosity (LOH) events spanning 1 to 40Mb and heterozygous events spanning 10 to 40Mb with a Total Copy Number (TCN) state between 3 and 9 were enriched in HRD samples (**Fig. 1*a-b***). In contrast, very large (>40Mb) heterozygous segments with TCN between 2 and 4 were enriched in HRP samples (**Fig. 1*a-b***). This finding suggests that very large diploid segments or regions that have undergone genome-doubling are enriched in HRP samples, in line with the observation that HRP samples are genomically stable and harbor relatively low copy number aberrations (18).

Based on these observations, we combined the mutational channels (**Methods**) into six genomic features: *(i)* genomic segments with LOH and sizes between 1 and 40 megabases (abbreviated as LOH:1-40Mb); *(ii)* deletions spanning at least 5bp at microhomologies (DEL.5.MH); *(iii)* heterozygous genomic segments with TCN between 3 and 9 and sizes between 10 and 40 megabases (3-9:HET:10-40Mb); *(iv)* C:G>G:C substitutions at 5’-Np<u>C</u>pT-3’ context (N[C>G]T); *(v)* C:G>T:A substitutions at 5’-Np<u>C</u>pG-3’ context (N[C>T]G); and *(vi)* heterozygous genomic segments with TCN between 2 and 4, and sizes above 40 megabases (2-4:HET:>40Mb). To evaluate if these genomic features are sufficient to distinguish HRD and HRP samples, we performed principal component analysis (PCA) using the training data. We observed a separation between HRD from HRP samples across the two principal components for both WGS (**Fig. 1*c***) and WES (**Fig. 1*d***) breast cancers.

Next, using the six genomic features, we trained a machine learning tool, HRProfiler, based on a linear kernel support vector machine. HRProfiler comprises WGS and WES models that were trained using 371 samples from the Sanger-WGS-Breast (13) and 672 samples from the TCGA-WES-Breast (17) datasets respectively (**Supplementary Fig. S1**). Ten-fold cross validation was conducted to determine the feature weights for the two trained models. As expected, features with positive weights (*i.e.*, LOH:1-40Mb, DEL.5.MH, 3-9:HET:10-40Mb, and N[C>G]T) were enriched in HRD samples, whereas features with negative weights (*i.e.*, N[C>T]G and 2-4:HET:>40Mb) were enriched in HRP samples (**Fig. 1*e***).

### Comparing HRD detection methods in WGS and WES breast cancers

In principle, two distinct approaches have been utilized to evaluate the performance of methods for detecting HRD. In their original publications, CHORD and HRDetect have relied on concordance between their predictions and prior HRD genomic annotations (13,14). This concordance can be quantified by area under the receiver operating characteristic curve (AUC) with both CHORD and HRDetect reporting AUCs above 0.90 for WGS cancers (13,14). However, this type of comparison requires a ground truth for HRD and HRP cancers which, in most cases, is not straightforward to derive. The second approach relies on comparing clinical endpoints for HRD and HRP predicted cancers in patients treated with either chemotherapy or PARPi. The advantage of this approach is that it could provide immediate clinical relevance. Unfortunately, such comparisons require the availability of well annotated clinico-genomics datasets which are currently limited especially at the whole-genome resolution. Here, we utilize both approaches to put HRProfiler in the context of previously developed methods.

To evaluate the performance of HRProfiler, SigMA, HRDetect, and CHORD in the context of HRD genomic ground truth annotations, we applied the four tools to an independent set of 237 whole-genome sequenced triple negative breast cancers (TNBCs) from the Sweden Cancerome Analysis Network – Breast project (SCAN-B; ClinicalTrials.gov identifier NCT02306096) (20) as well as to 71 held-out TCGA breast cancers which have been profiled using both whole-genome and whole-exome sequencing. Additionally, we applied the tools to an independent external WES dataset of 109 MSK-IMPACT breast cancers (21). All tools exhibited good AUC performance when applied to the WGS cancers (**Fig. 2*a-b*; Supplementary Fig. S2*a-b***) while HRProfiler outperformed HRDetect and SigMA for WES breast cancers (**Fig. 2*c-d*; Supplementary Fig. S2*c-d***). CHORD could not be applied to WES data. Importantly, HRProfiler was the only tool with AUCs above 0.90 across all WES and WGS breast cancer datasets (**Fig. 2**).

**Figure 2:**
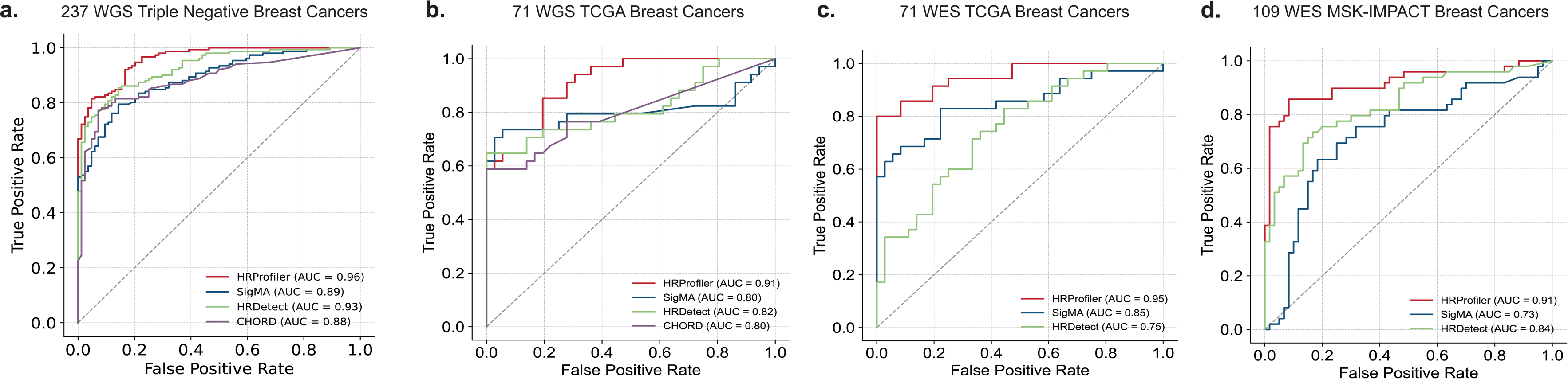
Performance of HRD tools on external validation datasets using HRD genomic ground truth annotations. Receiver operating characteristic curves (ROCs) were derived for HRProfiler, SigMA, HRDetect, and CHORD. Areas under the ROCs (AUCs) were calculated for each tool and shown in the legends of the respective panels. ***(a)*** ROCs for 237 whole-genome sequenced (WGS) triple negative breast cancers. ***(b)*** ROCs for 71 WGS TCGA breast cancers. ***(c)*** ROCs for 71 whole-exome sequenced (WES) breast cancers. ***(d)*** ROCs for 109 WES MSK-IMPACT breast cancers. No ROCs are shown for CHORD in panels *(c)* and *(d)* as the tool cannot be applied to WES data. In all plots, the x-axes reflect the false positive rates while the y-axes correspond to the true positive rates. Precision and recall curves for the same samples are provided in **Supplementary Figure S2**.

To evaluate the potential clinical utility of HRProfiler, SigMA, HRDetect, and CHORD in serving as predictive biomarkers for adjuvant chemotherapy treated breast cancers, we applied the tools to a subset of 145 whole-genome sequenced chemotherapy-treated TNBCs with information for interval disease-free survival (20). Additionally, the 145 TNBCs were down-sampled to whole-exomes (dWES) to further assess the ability of each tool to predict HRD robustly at both whole-genome and whole-exome resolutions. As previously reported (20), when applied to WGS breast cancers, HRDetect was able to identify 99 HRD samples which exhibited better survival when compared to the 46 HRP samples after adjusting for grade and age at diagnosis (hazard ratio [HR]=0.42; p-value=0.020; **Fig. 3*a***). However, the tool exhibited markedly worse sample stratification on the dWES data (HR=0.54; p-value=0.092) with 39 samples (26.9% of all examined TNBCs) being differently annotated when compared to the WGS data. CHORD’s performance on WGS samples was very similar to that of HRDetect (**Supplementary Fig. S3**), however, the tool cannot be applied to the dWES data. Applying SigMA to the 145 TNBCs did not result in a statistically significant separation for either the WGS breast cancers (p-value=0.068) or the dWES data (p-value=0.94; **Fig. 3*b***). In contrast, HRProfiler was able to better stratify breast cancers from both WGS (HR=0.40; p-value=0.021) and dWES data (HR=0.38; p-value=0.02; **Fig. 3*c***). Importantly, only 9 samples (6.2% of all examined TNBCs) were differently annotated by HRProfiler when the tool was applied to WGS and dWES data (**Fig. 3*c***). Lastly, partitioning the 145 TNBCs based on the presence of defects in *BRCA1/2* or the presence of HRD-associated signatures SBS3 or CN17 did not result in statistically significant separation (**Supplementary Fig. S4**). Nevertheless, stratifying the 145 TNBCs based on the presence of ID6 was able to separate the breast cancers, but captured 41 fewer HRD patients compared to HRProfiler (HR=0.48; p-value=0.04; **Supplementary Fig. S4**).

**Figure 3:**
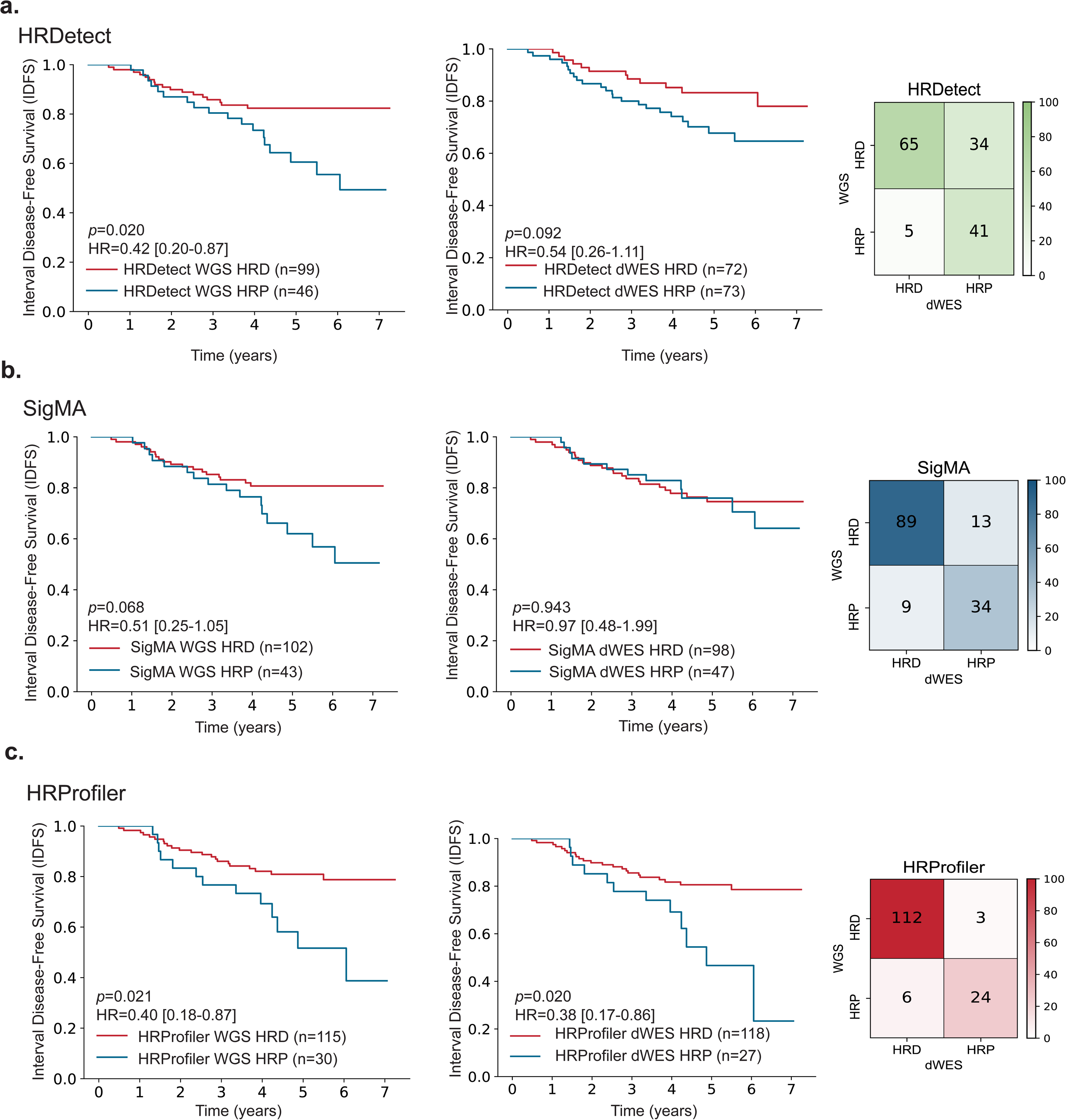
Predicting survival in breast cancers treated with chemotherapy by HRD tools. Kaplan-Meier curves and confusion matrices for samples predicted as HRD and HRP by ***(a)*** HRDetect, ***(b)*** SigMA, and ***(c)*** HRProfiler in 145 chemotherapy-treated triple negative breast cancers. In each panel, the left plot reflects the Kaplan-Meier curves for whole-genome sequenced breast cancers (WGS). The middle plot corresponds to the Kaplan-Meier curves for the same samples when down-sampled to whole-exomes (dWESs). The right plot contains a confusion matrix that provides a comparison of each tool’s HRD annotations from WGS and dWES data. The y-axes on all Kaplan-Meier curves reflect Interval Disease Free Survival (IDFS), and the x-axes correspond to time measured in years. Listed p-values and hazard ratios (HRs) are based on a Cox proportional hazards model after adjusting for age at diagnosis and tumor grade. 95% confidence intervals are provided for all HRs within the Kaplan-Meier plots. The performance of CHORD on WGS data, which was almost identical to the one of HRDetect, can be found in **Supplementary Figure S3**. Comparisons of the clinical utility of BRCA1/2 defects and HRD-associated signatures SBS3, CN17, and ID6 for the same patients are provided in **Supplementary Figure S4**.

### Comparing HRD detection methods in WES ovarian cancers

To determine if the breast cancer specific mutational features can be generalized to another HRD-associated cancer, we trained an ovarian-specific whole-exome model using 182 high-grade serous carcinoma from the TCGA-Ovarian-WES dataset (17) (**Supplementary Fig. S5*a***). As done for breast cancer, ten-fold cross validation was conducted for HRProfiler to determine the feature weights for the trained whole-exome model. Similar features to the ones observed in breast cancer were enriched in HRD and HRP ovarian cancers (**Supplementary Fig. S5*b***). To examine the performance of HRProfiler, SigMA, and HRDetect in the context of HRD genomic ground truth annotations for whole-exome sequenced ovarian cancer, we applied the three tools to 40 held-out TCGA ovarian samples as well as to an independent set of 50 MSK-IMPACT whole-exome sequenced ovarian cancers (21) (**Supplementary Fig. S6*a-b***). For both datasets, HRProfiler outperformed the other two approaches by consistently exhibiting AUCs above 0.90 (**Supplementary Fig. S6*a-b***).

To assess the clinical utility of HRProfiler, SigMA, and HRDetect to serve as predictors of clinical outcome in ovarian cancer, we examined the progression free survival for an independent set of 25 high-grade ovarian cancers from a phase Ib PARPi clinical trial of olaparib in combination with the PI3K inhibitor buparlisib (BKM120; ClinicalTrials.gov identifier NCT01623349) (22). HRProfiler’s annotations were able to separate PARPi treated samples based on progression free survival (HR=0.25; p-value=0.037; **Fig. 4**) with HRDetect also performing relatively well on these data (HR=0.32; p-value=0.056; **Fig. 4*b***). Moreover, partitioning the 25 PARPi-treated ovarian cancers based on the presence of any of the HRD-associated signatures SBS3, CN17, or ID6 did not lead to differences in survival endpoints (**Supplementary Fig. S7**). Lastly, annotating samples as HRD and HRP based on defects in *BRCA1/2* genes provided separation in progression free survival for the 25 PARPi-treated ovarian cancers (**Supplementary Fig. S7**).

**Figure 4:**
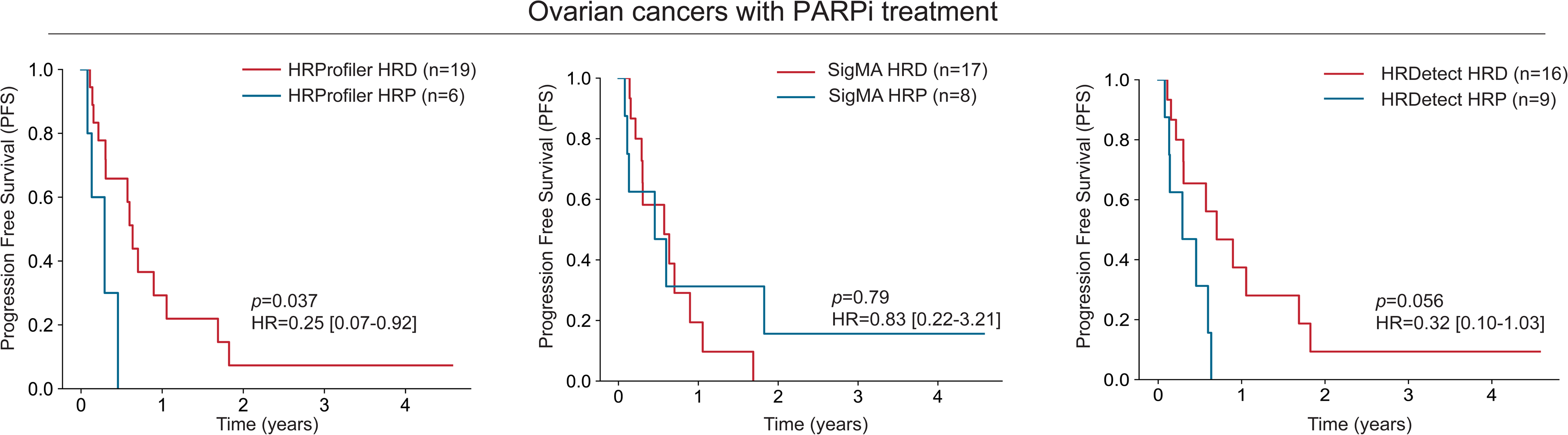
Predicting survival in ovarian cancers treated PARP inhibitor by HRD tools. Kaplan-Meier curves for progression free survival (PFS) across 25 PARP inhibitor treated patients with high-grade serous ovarian cancer. Patients are annotated as HRD or HRP based on the predictions from HRProfiler (*left panel*), SigMA (*middle panel*), and HRDetect (*right panel*). Listed p-values and hazard ratios (HRs) are based on a Cox proportional hazards model after adjusting for age at diagnosis and tumor stage. 95% confidence intervals are provided for all HRs within the Kaplan-Meier plots. Comparisons of the clinical utility of BRCA1/2 defects and HRD-associated signatures SBS3, CN17, and ID6 for the same patients are provided in **Supplementary Figure S7**.

## DISCUSSION

There is an increasing momentum in precision oncology towards more comprehensive genomic profiling to identify complex biomarkers like HRD as part of routine clinical care (23). With continuing advances in sequencing technologies and the corresponding exponential decrease in their cost, clinical whole-exome sequencing is becoming increasingly more prevalent (24–26). To harness the clinical utility of whole-exome sequencing for predicting HRD, we present a novel machine learning approach called HRProfiler that utilizes a minimal set of six genomic features to predict HRD across both whole-genome and whole-exome sequenced breast and ovarian cancers. Unlike existing methods that focus solely on mutation types enriched in HRD samples (13–15), HRProfiler incorporates small and large-scale mutational events enriched in both HRD and HRP cancers. HRProfiler also circumvents the need for genomic rearrangements and mutational signature extraction, which can be unreliable especially when using sparse datasets derived from whole-exome sequencing data (11).

HRProfiler demonstrated comparable performance to existing approaches when applied to whole-genome sequencing data and the tool surpassed other machine learning methods when applied to whole-exome sequenced cancers. The sub-optimal performance of HRDetect on whole-exome sequenced tumors is perhaps unsurprising given that HRDetect was developed for whole-genome sequenced breast cancers and the original publication noted a poor performance for whole-exome sequenced tumors (13). In contrast, despite its tailored design for whole-exome and targeted panel sequencing data, SigMA exhibited comparatively limited performance in our tests. Indeed, SigMA is a machine learning surrogate for detecting HRD-associated signature SBS3 and our results show that SBS3 alone is not a reliable predictor of survival even when detected by other tools. Similarly, other HRD-associated signatures, such as CN17 and ID6, did not provide consistent clinical separation for breast or ovarian cancers. Overall, these results indicate that the presence of an individual HRD-associated signature in a cancer sample does not necessarily indicate a clinically significant or an actionable event.

HRProfiler’s ability to separate HRD samples sensitive to treatment with PARP inhibitors from whole-exome sequencing data opens additional opportunities for broadening treatment options to a wider patient population. Given the non-tissue-specific nature of the HRD mutational footprint, our six mutational features can be refined in the future to predict HRD status in other HRD-associated cancers, including prostate and pancreatic cancers. Such an effort will ideally require large sets with well annotated clinico-genomics datasets for both cancer types, which, to the best of our knowledge, are currently not available.

Although we assessed HRProfiler’s performance using independent datasets encompassing 417 breast and 115 ovarian cancers, along with retrospective data from two clinical trials, we recognize the constraints posed by the use of relatively small sample sizes for some of the reported survival analyses. Future large-scale, independent, and purposefully designed clinical trials will be necessary to validate HRProfiler’s capacity to serve as a predictive and/or prognostic biomarker for routine clinical decision making. Notwithstanding, HRProfiler provides a crucial link in utilizing the molecular phenotypic changes of impaired DNA repair mechanisms for detecting homologous recombination deficiency in whole-exome sequenced cancers. Moreover, the tool provides a robust and consistent approach that allows detecting whole-exome sequenced cancers that are sensitive to PARP inhibitors.

## METHODS

### Data sources and pre-processing

In this study, previously published datasets were used for all feature engineering, model development, and validation for both whole-genome sequenced (WGS) and whole-exome sequenced (WES) breast and ovarian cancers.

For breast cancer, we downloaded CaVEman mutations and ASCAT allele-specific copy number for 560 Sanger breast cancers (10) from: ftp://ftp.sanger.ac.uk/pub/cancer/Nik-ZainalEtAl-560BreastGenomes/. Additional WGS breast cancer datasets used in this study included the 237 Triple Negative Breast (TNBC) samples from the SCAN-B clinical trial (20). CaVEman somatic mutations and ASCAT copy number for the 237 TNBC samples were downloaded from: https://data.mendeley.com/datasets/2mn4ctdpxp/. For the breast cancer WGS dataset from the Pan-Cancer Analysis of Whole Genomes project, consensus somatic mutations and copy number calls were downloaded from the International Cancer Genome Consortium’s data portal: https://dcc.icgc.org/releases/PCAWG. For The Cancer Genome Atlas (TCGA) breast cancer WES dataset, the catalogues of somatic mutations and sequencing data were downloaded from the genomics data commons (https://portal.gdc.cancer.gov/) portal and allele-specific whole-exome copy number calls were derived using ASCAT: https://github.com/VanLoo-lab/ascat. For the WES MSK-IMPACT breast cancers, 109 whole-exome sequenced breast cancers were downloaded from dbGaP (accession number: phs001783.v1.p1) and processed using an ensemble variant calling pipeline: https://github.com/AlexandrovLab/EnsembleVariantCallingPipeline

For ovarian cancer, the WES derived catalogues of somatic mutations and sequencing data from TCGA were downloaded from the genomics data commons portal, and allele-specific whole-exome copy number calls were derived using ASCAT. For the ovarian cancer WES MSK-IMPACT dataset, 50 whole-exome sequenced ovarian cancers were downloaded from dbGaP (accession number: phs001783.v1.p1) and processed using the same ensemble variant calling pipeline as the one utilized for breast cancer. Lastly, we downloaded the 25 PARPi treated high-grade ovarian cancers from dbGaP (accession number: phs003019) and processed these data using the ensemble variant calling pipeline.

### Feature engineering for predicting HRD

As previously done (13,14,18), a sample with an HRD score of at least 42 for breast cancer (5) and 63 for ovarian cancer (27) or one harboring germline/somatic alterations in *BRCA1* or *BRCA2* was annotated as homologous recombination deficient (HRD) for all training purposes. All other samples were annotated as homologous recombination proficient (HRP). To identify significantly enriched features in HRD and HRP samples, we generated the average mutational profiles based on proportions across the 96 substitution, 83 indel, and 48 copy number mutational contexts. To determine differences in channels at every resolution, we performed Fisher’s exact tests to evaluate if there is any statistically significant difference in the average proportion of a given channel between HRD and HRP samples. Significant channels were identified for all types of mutational contexts if their absolute log_2_ fold-change (FC) was greater than 0.75 for WGS samples and 0.25 for WES samples, and their −log_10_(FDR adjusted p-value) was greater than 3. Similar workflow was adopted for both whole-genome and whole-exome samples and only channels significantly enriched across both WGS and WES were considered for the feature engineering process. At the single base resolution, A[C>T]G, C[C>T]G, G[C>T]G, and T[C>T]G channels were consistently enriched across HRP samples in both whole-genome and whole-exome datasets. Due to the overlapping/similar mutational context, these four channels were combined into a single feature termed N[C>T]G, where N represents any of the four nucleotide bases (A, C, T, or G). Similarly, A[C>G]T, C[C>G]T, G[C>G]T, and T[C>G]T were channels consistently enriched in HRD samples and were combined into a single feature N[C>G]T. At the indel resolution, 5:Del:M:1, 5:Del:M:2, 5:Del:M:3, 5:Del:M:4, and 5:Del:M:5 were significantly enriched channels in HRD samples that represent varying lengths of microhomology sequences at relatively large deletion sites where the length of the deletion is at least 5 base pairs long. These indel channels were combined into a single feature: DEL.5.MH, where DEL.5 presents deletions of length at least 5 bp and MH represent microhomology sequences. At the copy number resolution, multiple significant Loss of Heterozygosity (LOH) events were identified. These events represented LOH segments of at least 1 Mb, where majority of the segment sizes ranged between 1 and 40Mb. These were combined into a single feature LOH:1-40Mb. A similar approach was applied to aggregate significant copy number channels for diploid/genome-doubled copy number segments into a single feature 2-4:HET:>40Mb that accounts for segments with a total copy number state between 2-4 and sizes of at least 40Mb. Lastly, significant copy number channels for amplification events were combined into a single feature: 3-9:HET:10-40Mb, where 3-9 represents the segments with a total copy number state of at least 3 and segment sizes between 10 to 40Mb.

### Training and comparing HRD detection methods in WGS cancers

To train a model for predicting HRD at WGS resolution, we used samples from the 560 Breast dataset. Only 371/560 samples that were labelled as evaluated in the HRDetect publication (13) were considered. The six features derived from the feature engineering step were extracted from the 371 samples and were normalized using StandardScaler in python’s sklearn package. The training was based on 371 breast samples, comprising 131 HRD and 240 HRP samples, and used a linear kernel support vector machine with L2 regularization. Next, 10-fold cross validation was conducted to tune for hyper-parameters and obtain feature weights from the model. To test the model’s performance, we predicted HRD probabilities for 71 WGS TCGA breast samples that were sequenced at both whole-genome and whole-exome resolutions. Samples with an HRD probability at least 0.50 were considered as HRD. To validate the model on an external dataset, we predicted HRD probabilities for 237 Triple Negative Breast (TNBC) samples and evaluated its performance against the ground truth. The performance of the model was assessed using machine learning metrics such as sensitivity, precision, and F_1_ score. To compare the performance of HRProfiler with other tools, HRD annotations were determined for the 237 TNBC samples using HRDetect, CHORD, and SigMA.

### Training and comparing HRD detection methods in WES cancers

To train a breast cancer specific model for predicting HRD at WES resolution, we used samples from TCGA breast cancer dataset. Only 743 samples that had HRD annotations were used for both training and testing. The six features derived from the feature engineering step were extracted as proportions, except for DEL.5.MH, which was extracted as absolute counts. All features were normalized using StandardScaler in python’s sklearn package. The training was based on 672 breast samples that included 156 HRD and 516 HRP samples. Next, 10-fold cross validation was conducted to tune for hyper-parameters and obtain feature weights from the model. The model’s performance was tested on the held-out 71 breast samples that were previously sequenced at both whole-genome and whole-exome resolution. Samples with an HRD probability at least 0.50 were considered as HRD. To validate the model on an external dataset, we predicted HRD probabilities for 109 MSK-IMPACT breast cancer whole-exome sequenced samples and evaluated the model’s performance against the ground truth. The performance of the model was assessed using conventional machine learning metrics such as sensitivity, precision, and F_1_ score. The WES model was also applied to the down-sampled 237 TNBC samples. The whole-exome features for the 237 TNBC samples were derived by down-sampling the ASCAT copy number calls to segments that spanned the exonic regions. The mutation and indel calls were down-sampled to whole-exome resolution using SigProfiler (28). To compare the performance of HRProfiler with other tools, HRD probabilities were also determined for SigMA and HRDetect.

To train an ovarian-specific model for predicting HRD at WES resolution, we used samples from the TCGA ovarian dataset. Only 228 samples that had HRD annotations were used for both training and testing. Analogous to training HRProfiler for WES breast cancers, the six features derived from the feature engineering step were extracted as proportions, except for DEL.5.MH, which was extracted as absolute counts. All features were normalized using StandardScaler in the python sklearn package. The training was based on 182 ovarian cancers that comprised of 82 HRD and 100 HRP samples. Next, 10-fold cross validation was conducted to tune for hyper-parameters and obtain feature weights from the model. The model’s performance was tested on the 39 ovarian cancer that were sequenced at whole-exome resolution. Samples with an HRD probability at least 0.50 were considered as HRD. To validate the model on an external dataset, we predicted HRD probabilities for 50 MSK-IMPACT whole-exome sequenced ovarian cancers and evaluated the model’s performance against the ground truth. The performance of the model was assessed using conventional machine learning metrics such as sensitivity, precision, and F_1_ score. To compare the performance of HRProfiler with other tools, HRD annotations were determined for the same samples by HRDetect and SigMA using the default breast WGS and ovarian WES pre-trained models, respectively.

### Deriving HRD status based on HRD-associated signatures, genes, and tools

Germline and somatic mutations for *BRCA1* and *BRCA2* and, when available, gene expression and promoter methylation changes in *BRCA1* and *BRCA2* were incorporated for the *BRCA1/2* annotations. Specifically, for TCGA breast cancers, the *BRCA1/2* annotations were derived from Polak *et al.* (29). Conversely, for TCGA ovarian cancers, these annotations were derived from Steele *et al.* (12). For all other datasets, BRCA1/2 annotations were derived from their respective publications.

SigprofilerAssigment (v0.1.2) was used to determine the presence of HRD-associated signatures SBS3, ID6, and CN17 (30) using the Catalogue Of Somatic Mutations In Cancer (COSMICv3.4) reference signatures. A sample was classified as HRD positive for a given HRD signature, if it had at least one mutational event attributed to that signature.

HRDetect was run using the Signature.tools.lib (v2.3.0) package in R, available at https://github.com/Nik-Zainal-Group/signature.tools.lib. The default HRD probability threshold of 0.70 was employed for predicting HRD status for WGS samples. To execute HRDetect on WES data, we utilized the pre-trained WGS model for prediction. The rearrangement signatures RS3 and RS5, which cannot be derived from WES data, were set to zero, and the default probability threshold of 0.70 was applied for classifying whole-exome sequenced cancers as HRD.

CHORD was run using the extractSigsChord function installed from GitHub: https://github.com/UMCUGenetics/CHORD/. It was executed using default settings, and a probability threshold of 0.50 was applied for classifying samples as HRD.

SigMA (v2.0) was downloaded from GitHub: https://github.com/parklab/SigMA/archive/refs/tags/2.0.tar.gz and it was run using the run function for signature 3 (also known as SBS3) prediction. For WGS breast datasets, we used the following parameters when running SigMA: data=’wgs’, do_assign=T, do_mva=T, tumor_type=’breast’, and catalog_name=’cosmic_v3p2_inhouse’, and we utilized SigMA strict predictions (pass_mva_strict) for our analysis. When running SigMA on WES datasets, we followed the same procedure as for WGS datasets, except for the data and tumor_type parameters. For predicting signature 3 status for TCGA datasets, the data parameter was set to ‘tcga_mc3’, otherwise, it was set to ‘seqcap’ for all other WES and down-sampled WES datasets. The tumor_type parameter was set to ‘breast’ for breast and ‘ovary’ for ovarian whole-exome sequencing data.

### Survival analysis

The survival analysis was conducted using the KaplanMeierFitter and CoxPHFitter function from the lifelines package in python (31). Interval disease free survival was used to evaluate patients treated with chemotherapy from the 237 TNBC dataset. Progression free survival endpoint was used to evaluate the survival trends for 25 high-grade ovarian cancer patients treated with PARP inhibitor. P-values and hazard ratios listed in the Kaplan Meier plots are based on the p-values derived from the Cox proportional hazards (coxph) model adjusted by dichotomized age of diagnosis (below and above 50 years old) as well as tumor stage or grade.

### Statistics

All statistical analysis were conducted in python using the scikit-learn package. All p-values were corrected for multiple hypothesis testing using Benjamini-Hochberg procedure, where applicable.

## Data Availability

All data used in this study were previously generated by others and are publicly available.

https://www.ncbi.nlm.nih.gov/projects/gap/cgi-bin/study.cgi?study_id=phs001783.v6.p1

https://www.ncbi.nlm.nih.gov/projects/gap/cgi-bin/study.cgi?study_id=phs003019.v1.p1

ftp://ftp.sanger.ac.uk/pub/cancer/Nik-ZainalEtAl-560BreastGenomes/

https://data.mendeley.com/datasets/2mn4ctdpxp/

https://dcc.icgc.org/releases/PCAWG

## Availability of data and materials

HRProfiler is an open-source tool, and it is freely available for academic use as a python package at https://github.com/AlexandrovLab/HRProfiler. The pre-trained models for whole-genome and whole-exome sequenced breast and ovarian cancers are provided as part of the tool.

## Funding and acknowledgements

This work was supported by the US National Institute of Health grants R01ES030993-01A1, U01DE033345, R01ES032547-01, and R01CA269919-01 to LBA as well as LBA’s Packard Fellowship for Science and Engineering and Cancer Research UK Grand Challenge Award C98/A24032. The research in this grant was also supported by a Curebound Targeted grant to LBA and RRM. The computational analyses reported in this manuscript have utilized the Triton Shared Computing Cluster at the San Diego Supercomputer Center of UC San Diego. The funders had no roles in study design, data collection and analysis, decision to publish, or preparation of the manuscript.

## Authors’ contributions

AA and LBA designed the overall study. AA performed all analyses with help from CDS, ENB, AK, AF, RRM, NP, and LBA. CDS and AK assisted in the copy number analysis and the feature engineering process. AF contributed to testing the tool’s functionality. ENB, RRM, NP, and LBA assisted in the interpretation and analysis of the survival and clinical associations. AA and LBA wrote the manuscript with help and input from all other authors. All authors read and approved the final manuscript.

## REFERENCES

1. Konstantinopoulos PA, Ceccaldi R, Shapiro GI, D’Andrea AD. Homologous Recombination Deficiency: Exploiting the Fundamental Vulnerability of Ovarian Cancer. Cancer Discov 2015;5(11):1137–54 doi 10.1158/2159-8290.CD-15-0714.

2. Moore K, Colombo N, Scambia G, Kim BG, Oaknin A, Friedlander M, et al. Maintenance Olaparib in Patients with Newly Diagnosed Advanced Ovarian Cancer. N Engl J Med 2018;379(26):2495–505 doi 10.1056/NEJMoa1810858.

3. Tutt A, Tovey H, Cheang MCU, Kernaghan S, Kilburn L, Gazinska P, et al. Carboplatin in BRCA1/2-mutated and triple-negative breast cancer BRCAness subgroups: the TNT Trial. Nat Med 2018;24(5):628–37 doi 10.1038/s41591-018-0009-7.

4. Birkbak NJ, Wang ZC, Kim JY, Eklund AC, Li Q, Tian R, et al. Telomeric allelic imbalance indicates defective DNA repair and sensitivity to DNA-damaging agents. Cancer Discov 2012;2(4):366–75 doi 10.1158/2159-8290.CD-11-0206.

5. Telli ML, Timms KM, Reid J, Hennessy B, Mills GB, Jensen KC, et al. Homologous Recombination Deficiency (HRD) Score Predicts Response to Platinum-Containing Neoadjuvant Chemotherapy in Patients with Triple-Negative Breast Cancer. Clin Cancer Res 2016;22(15):3764–73 doi 10.1158/1078-0432.CCR-15-2477.

6. Abkevich V, Timms KM, Hennessy BT, Potter J, Carey MS, Meyer LA, et al. Patterns of genomic loss of heterozygosity predict homologous recombination repair defects in epithelial ovarian cancer. Br J Cancer 2012;107(10):1776–82 doi 10.1038/bjc.2012.451.

7. Zamborszky J, Szikriszt B, Gervai JZ, Pipek O, Poti A, Krzystanek M, et al. Loss of BRCA1 or BRCA2 markedly increases the rate of base substitution mutagenesis and has distinct effects on genomic deletions. Oncogene 2017;36(35):5085–6 doi 10.1038/onc.2017.213.

8. Petljak M, Alexandrov LB, Brammeld JS, Price S, Wedge DC, Grossmann S, et al. Characterizing Mutational Signatures in Human Cancer Cell Lines Reveals Episodic APOBEC Mutagenesis. Cell 2019;176(6):1282–94 e20 doi 10.1016/j.cell.2019.02.012.

9. Alexandrov LB, Nik-Zainal S, Wedge DC, Aparicio SA, Behjati S, Biankin AV, et al. Signatures of mutational processes in human cancer. Nature 2013;500(7463):415-21 doi 10.1038/nature12477.

10. Nik-Zainal S, Davies H, Staaf J, Ramakrishna M, Glodzik D, Zou X, et al. Landscape of somatic mutations in 560 breast cancer whole-genome sequences. Nature 2016;534(7605):47-54 doi 10.1038/nature17676.

11. Alexandrov LB, Kim J, Haradhvala NJ, Huang MN, Tian Ng AW, Wu Y, et al. The repertoire of mutational signatures in human cancer. Nature 2020;578(7793):94-101 doi 10.1038/s41586-020-1943-3.

12. Steele CD, Abbasi A, Islam SMA, Bowes AL, Khandekar A, Haase K, et al. Signatures of copy number alterations in human cancer. Nature 2022;606(7916):984-91 doi 10.1038/s41586-022-04738-6.

13. Davies H, Glodzik D, Morganella S, Yates LR, Staaf J, Zou X, et al. HRDetect is a predictor of BRCA1 and BRCA2 deficiency based on mutational signatures. Nat Med 2017;23(4):517–25 doi 10.1038/nm.4292.

14. Nguyen L, J WMM, Van Hoeck A, Cuppen E. Pan-cancer landscape of homologous recombination deficiency. Nat Commun 2020;11(1):5584 doi 10.1038/s41467-020-19406-4.

15. Gulhan DC, Lee JJ, Melloni GEM, Cortes-Ciriano I, Park PJ. Detecting the mutational signature of homologous recombination deficiency in clinical samples. Nat Genet 2019;51(5):912–9 doi 10.1038/s41588-019-0390-2.

16. Zehir A, Benayed R, Shah RH, Syed A, Middha S, Kim HR, et al. Mutational landscape of metastatic cancer revealed from prospective clinical sequencing of 10,000 patients. Nat Med 2017;23(6):703–13 doi 10.1038/nm.4333.

17. Gao GF, Parker JS, Reynolds SM, Silva TC, Wang LB, Zhou W, et al. Before and After: Comparison of Legacy and Harmonized TCGA Genomic Data Commons’ Data. Cell Syst 2019;9(1):24–34 e10 doi 10.1016/j.cels.2019.06.006.

18. Marquard AM, Eklund AC, Joshi T, Krzystanek M, Favero F, Wang ZC, et al. Pan-cancer analysis of genomic scar signatures associated with homologous recombination deficiency suggests novel indications for existing cancer drugs. Biomark Res 2015;3:9 doi 10.1186/s40364-015-0033-4.

19. Pettitt SJ, Frankum JR, Punta M, Lise S, Alexander J, Chen Y, et al. Clinical brca1/2 reversion analysis identifies hotspot mutations and predicted neoantigens associated with therapy resistance. Cancer Discovery 2020;10(10):1475–88 doi 10.1158/2159-8290.CD-19-1485.

20. Staaf J, Glodzik D, Bosch A, Vallon-Christersson J, Reutersward C, Hakkinen J, et al. Whole-genome sequencing of triple-negative breast cancers in a population-based clinical study. Nat Med 2019;25(10):1526–33 doi 10.1038/s41591-019-0582-4.

21. Jonsson P, Bandlamudi C, Cheng ML, Srinivasan P, Chavan SS, Friedman ND, et al. Tumour lineage shapes BRCA-mediated phenotypes. Nature 2019;571(7766):576-9 doi 10.1038/s41586-019-1382-1.

22. Batalini F, Gulhan DC, Mao V, Tran A, Polak M, Xiong N, et al. Mutational Signature 3 Detected from Clinical Panel Sequencing is Associated with Responses to Olaparib in Breast and Ovarian Cancers. Clin Cancer Res 2022;28(21):4714–23 doi 10.1158/1078-0432.CCR-22-0749.

23. Menzel M, Ossowski S, Kral S, Metzger P, Horak P, Marienfeld R, et al. Multicentric pilot study to standardize clinical whole exome sequencing (WES) for cancer patients. NPJ Precis Oncol 2023;7(1):106 doi 10.1038/s41698-023-00457-x.

24. Van Allen EM, Wagle N, Stojanov P, Perrin DL, Cibulskis K, Marlow S, et al. Whole-exome sequencing and clinical interpretation of formalin-fixed, paraffin-embedded tumor samples to guide precision cancer medicine. Nat Med 2014;20(6):682–8 doi 10.1038/nm.3559.

25. Horak P, Heining C, Kreutzfeldt S, Hutter B, Mock A, Hullein J, et al. Comprehensive Genomic and Transcriptomic Analysis for Guiding Therapeutic Decisions in Patients with Rare Cancers. Cancer Discov 2021;11(11):2780–95 doi 10.1158/2159-8290.CD-21-0126.

26. Niguidula N, Alamillo C, Shahmirzadi Mowlavi L, Powis Z, Cohen JS, Farwell Hagman KD. Clinical whole-exome sequencing results impact medical management. Mol Genet Genomic Med 2018;6(6):1068–78 doi 10.1002/mgg3.484.

27. Takaya H, Nakai H, Takamatsu S, Mandai M, Matsumura N. Homologous recombination deficiency status-based classification of high-grade serous ovarian carcinoma. Sci Rep 2020;10(1):2757 doi 10.1038/s41598-020-59671-3.

28. Bergstrom EN, Huang MN, Mahto U, Barnes M, Stratton MR, Rozen SG, et al. SigProfilerMatrixGenerator: a tool for visualizing and exploring patterns of small mutational events. BMC Genomics 2019;20(1):685 doi 10.1186/s12864-019-6041-2.

29. Polak P, Kim J, Braunstein LZ, Karlic R, Haradhavala NJ, Tiao G, et al. A mutational signature reveals alterations underlying deficient homologous recombination repair in breast cancer. Nat Genet 2017;49(10):1476–86 doi 10.1038/ng.3934.

30. Diaz-Gay M, Vangara R, Barnes M, Wang X, Islam SMA, Vermes I, et al. Assigning mutational signatures to individual samples and individual somatic mutations with SigProfilerAssignment. Bioinformatics 2023;39(12) doi 10.1093/bioinformatics/btad756.

31. Davidson-Pilon C. lifelines: survival analysis in Python. Journal of Open Source Software 2019;4(40) doi 10.21105/joss.01317.

