## Supplementary Figures S1-S7 for "Detecting HRD in whole-genome and whole-exome sequenced breast and ovarian cancers"

This supplementary data has been provided by the authors to give readers additional information about their work.

### Supplementary Data

**Section S1: Supplementary Figures S1-S7……………………………….……………3**

Figure S1: Datasets used for training, testing, and validating HRProfiler in breast cancers ……………………..……………………...………………………………………..…..…………..3

Figure S2: Precision and recall of HRD tools on breast validation datasets using HRD genomic ground truth annotations ………………………...................................................………....4

Figure S3: Evaluating CHORD for predicting survival to chemotherapy in whole-genome sequenced breast cancers…………………………...................………..……...............….5

Figure S4: Evaluating the presence of defects in BRCA1/2 or HRD-associated signatures for predicting survival in chemotherapy-treated breast cancers……………………………......…..6

Figure S5: Datasets used for training, testing, and validating HRProfiler in ovarian cancers ……………………..……………………...…………………..……….........................................…....7

Figure S6: Performance of HRD tools on external ovarian validation datasets using HRD genomic ground truth annotations.....................................................................................................8

Figure S7: Evaluating the presence of defects in BRCA1/2 or HRD-associated signatures for predicting survival in PARP inhibitor treated ovarian cancer...……………………………..........9

### Section S1: Supplementary Figures

**Figure S1: Datasets used for training, testing, and validating HRProfiler in breast cancer.** Schematic outline of the workflow for training, testing, and validating HRProfiler, a support vector machine model for detecting homologous recombination deficient (HRD) and homologous recombination proficient (HRP) breast cancers from whole-genome sequenced (WGS) and whole-exome sequenced (WES) data. The model was trained based on 6 genomic features, encompassing, single base substitutions (SBS), small insertions and deletions (ID), and copy-number alternations (CN). Training and testing data included samples from The Cancer Genome Atlas (TCGA), Sanger institute, and Pan-Cancer Analysis of Whole Genomes (PCAWG) study. Validation datasets include the independent Triple Negative Breast (TNBC) and the Memorial Sloan Kettering Cancer Center's Integrated Mutation Profiling of Actionable Cancer Targets (MSK-IMPACT) data.

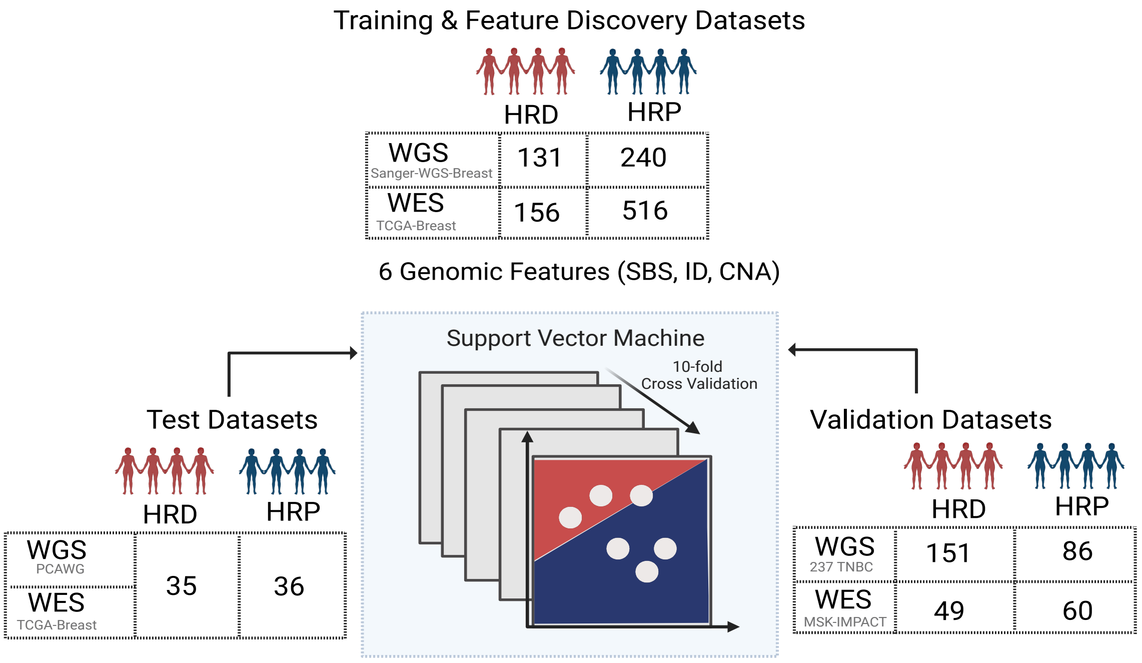

**Figure S2: Precision and recall of HRD tools on breast validation datasets using HRD genomic ground truth annotations.** Precision and recall (PR) curves were calculated for HRProfiler, SigMA, HRDetect, and CHORD. ***(a)*** PR curves for 237 whole-genome sequenced (WGS) triple negative breast cancers. ***(b)*** PR curves for 71 WGS breast cancers from The Cancer Genome Atlas (TCGA) project. ***(c)*** PR curves for 71 whole-exome sequenced (WES) breast cancers. ***(d)*** PR curves for 109 MSK-IMPACT WES breast cancers. No PR curves are shown for CHORD in panels *(c)* and *(d)* as the tool cannot be applied to WES data. The $F_1$ scores, *i.e.*, the harmonic mean of precision and recall, are shown for each tool within the respective panel's legend.

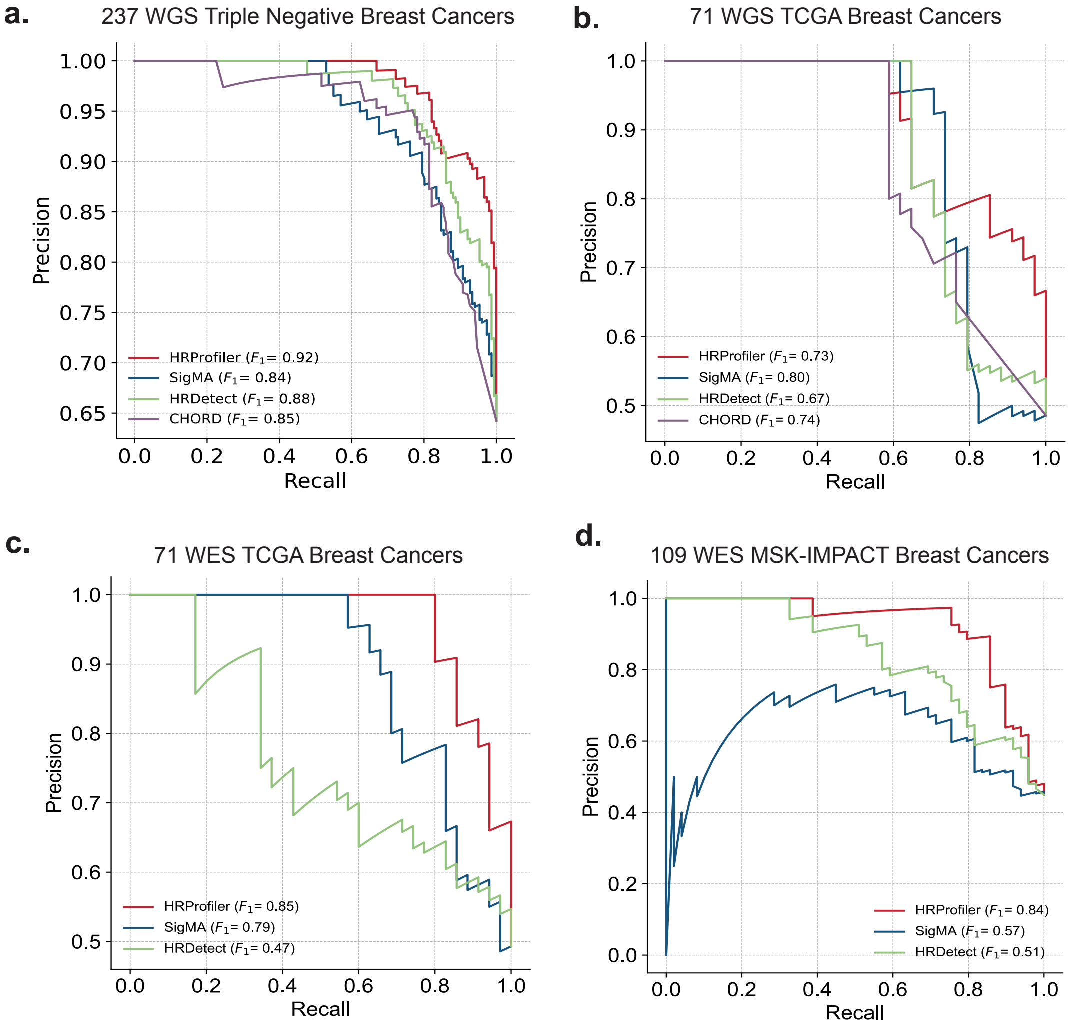

**Figure S3: Evaluating CHORD for predicting survival to chemotherapy in whole-genome sequenced breast cancers.** All presented results are for 145 chemotherapy-treated whole-genome sequenced (WGS) triple negative breast cancers. ***(a)*** Kaplan-Meier curves for 131 breast cancers annotated as HRD and HRP by CHORD. Note that 14/145 samples were classified as undetermined by CHORD and these samples were excluded from the survival analysis. The y-axis of the Kaplan-Meier curves reflects Interval Disease Free Survival (IDFS), and the x-axis corresponds to time measured in years. The p-value and hazard ratio (HR) are based on a Cox proportional hazards model after adjusting for age and tumor grade. An 95% confidence interval is provided for the HR within the Kaplan-Meier plot. ***(b)*** Confusion matrix comparing the HRD and HRP annotations between CHORD and HRDetect.

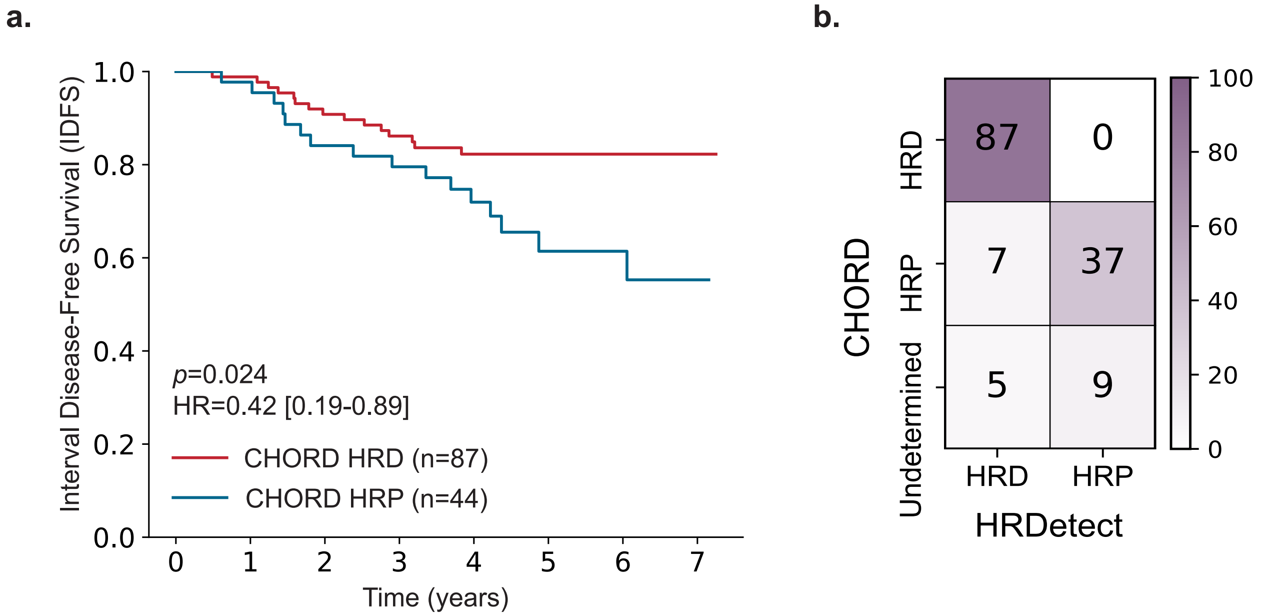

**Figure S4: Evaluating the presence of defects in *BRCA1/2* or HRD-associated signatures for predicting survival in chemotherapy-treated breast cancers.** All presented results are for 145 chemotherapy-treated triple negative breast cancers down-sampled to whole-exomes (dWES). Kaplan-Meier curves and hazard ratios (HRs) for samples annotated as HRD and HRP by either a defect in *BRCA1/2* or by the presence of HRD-associated signatures SBS3, CN17, or ID6. The p-values and HRs are based on a Cox proportional hazards model after adjusting for age and tumor grade. 95% confidence intervals are provided for the HRs within the Kaplan-Meier plots. The y-axes on all Kaplan-Meier curves reflect Interval Disease Free Survival (IDFS), and the x-axes correspond to time measured in years.

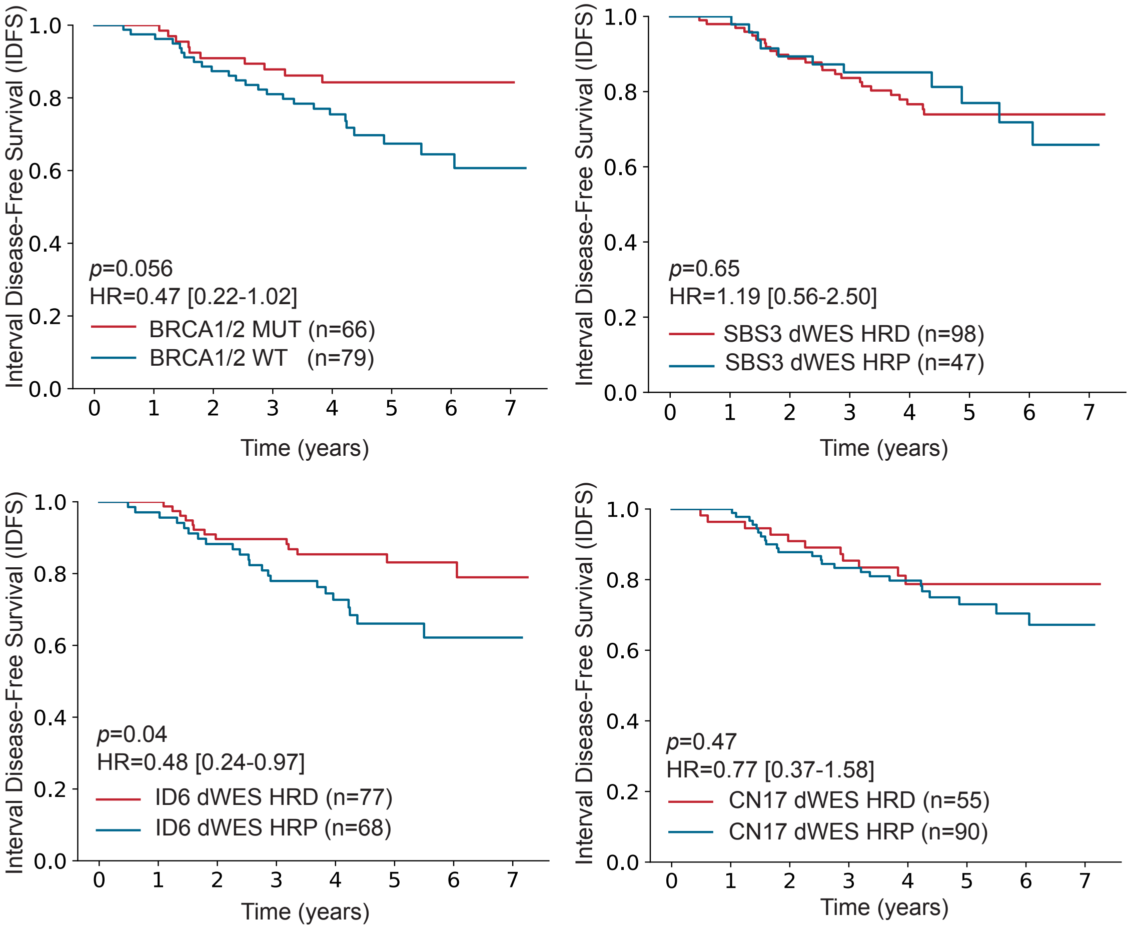

**Figure S5: Datasets and features used for training, testing, and validating HRProfiler in ovarian cancer *(a)*** Schematic outline of the workflow for training, testing, and validating HRProfiler, a support vector machine model for detecting homologous recombination deficient (HRD) and homologous recombination proficient (HRP) ovarian cancers from whole-exome sequenced data. The model was trained based on 6 genomic features, encompassing, single base substitutions (SBS), small insertions and deletions (ID), and copy-number alternations (CN). Training and testing data included samples from The Cancer Genome Atlas (TCGA) project. Validation datasets include the independent Memorial Sloan Kettering Cancer Center's Integrated Mutation Profiling of Actionable Cancer Targets (MSK-IMPACT) dataset and samples from a phase Ib trial of the PARP inhibitor olaparib in combination with the PI3K inhibitor (BKM120 cohort). ***(b)*** The average 10-fold cross validation weights of the six features derived from WES ovarian training dataset using a linear-kernel support vector machine. Positive weights reflect features predictive for HRD samples, while negative weights correspond to features predictive for HRP samples.

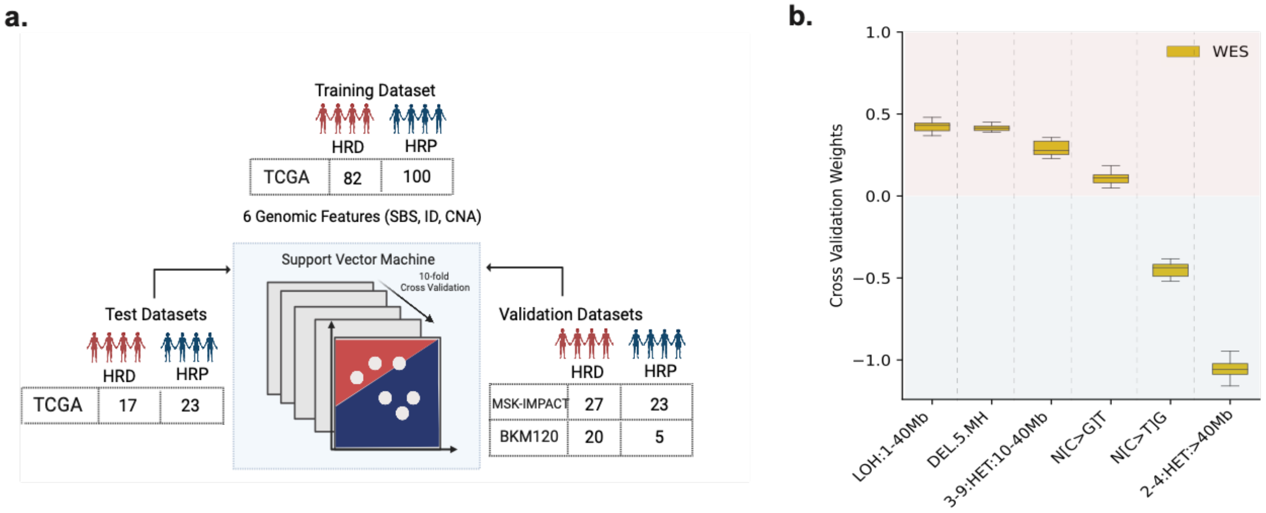

**Figure S6: Performance of HRD tools on external ovarian validation datasets using HRD genomic ground truth annotations.** ***(a)*** Receiver operating characteristic (ROC) curves and ***(b)*** precision and recall curves were calculated for HRProfiler, SigMA, and HRDetect on a held-out test dataset of 40 whole-exome sequenced (WES) ovarian samples from The Cancer Genome Atlas (TCGA) project and on an external validation dataset of 50 WES MSK-IMPACT ovarian cancer. The areas under the ROC (AUCs) as well as the $F_1$ scores, *i.e.*, the harmonic mean of precision and recall, are shown for each tool within the respective legend of each panel.

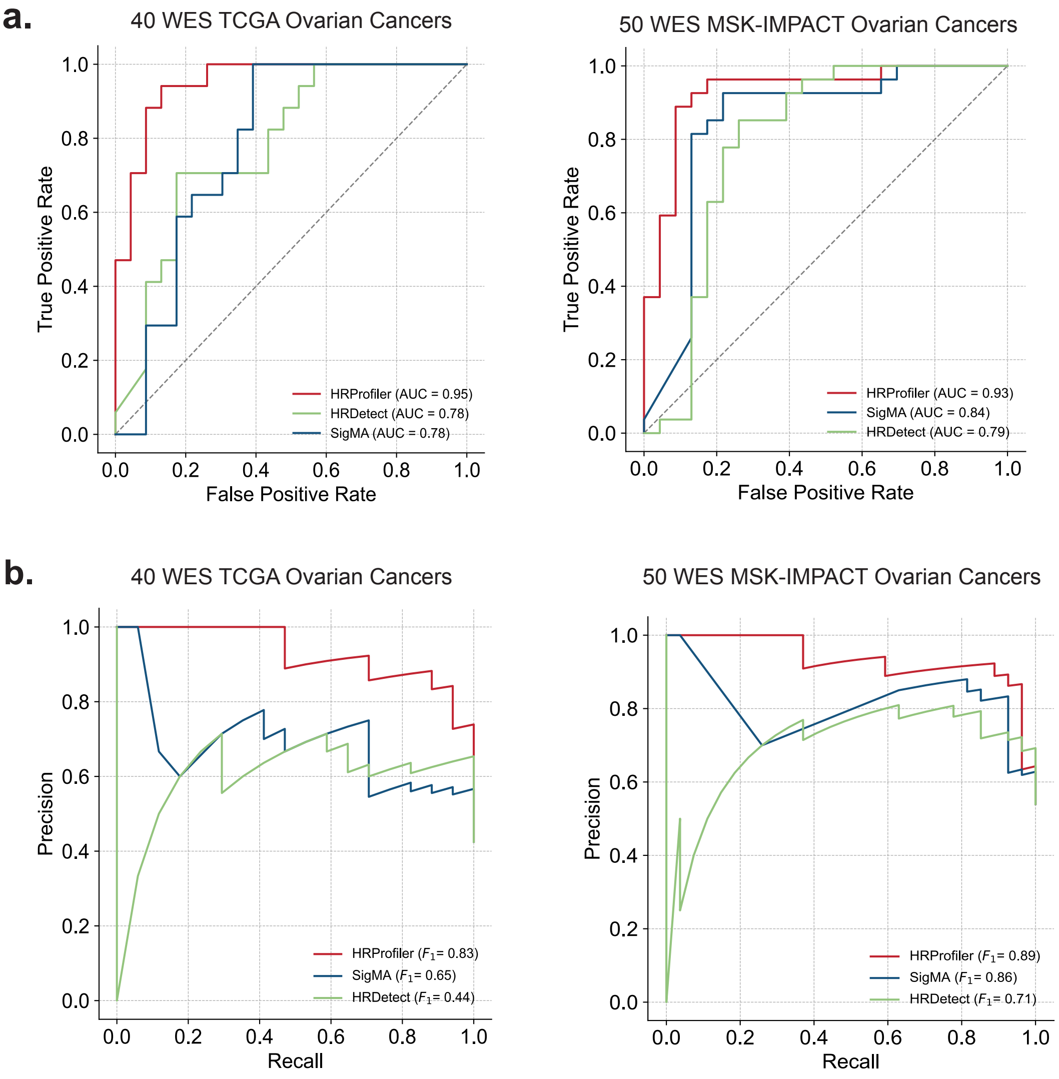

**Figure S7: Evaluating the presence of defects in *BRCA1/2* or HRD-associated signatures for predicting survival in PARP inhibitor treated ovarian cancers.** Progression free survival (PFS) across 25 PARPi treated ovarian cancers stratified based on presence of ***(a)*** BRCA1/2 mutations, ***(b)*** SBS3, ***(c)*** CN17, or ***(d)*** ID6.Listed p-values and hazard ratios (HRs) are based on a Cox proportional hazards model after adjusting for age at diagnosis and tumor stage. 95% confidence intervals are provided for all HRs within the Kaplan-Meier plots.

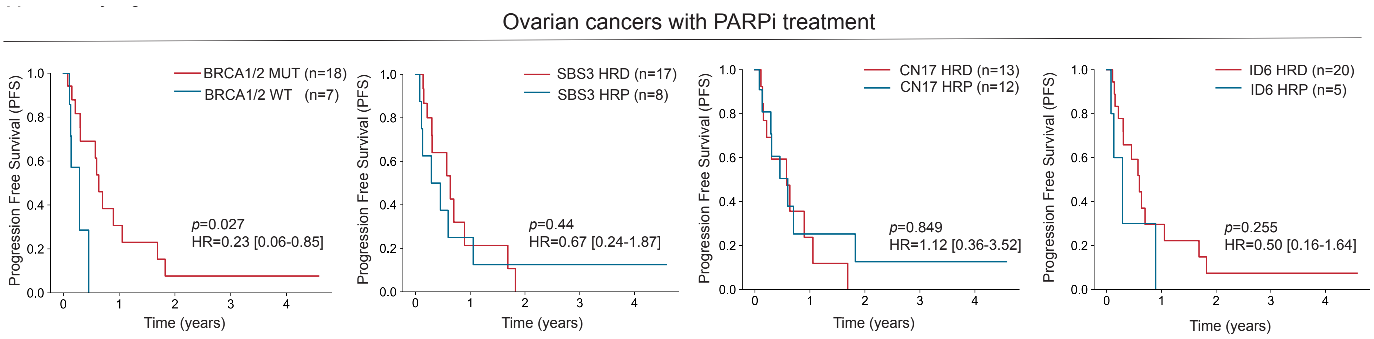
